# Efficacy of Dapagliflozin on Recurrence after Catheter Ablation for Atrial Fibrillation: the Statistical Analysis Plan of the DARE-AF Trial

**DOI:** 10.1101/2025.06.19.25329830

**Authors:** Zixu Zhao, Zejun Yang, Chi Wang, Chao Jiang, Jianzeng Dong, Changsheng Ma

**Author notes:** Correspondence authors: Chao Jiang Department of Cardiology, Beijing Anzhen Hospital, Capital Medical University, No. 2, Anzhen Road, Chao Yang District, Beijing, China, 100029; E Changsheng Ma Department of Cardiology, Beijing Anzhen Hospital, Capital Medical University, No. 2, Anzhen Road, Chao Yang District, Beijing, China, 100029; E. The authors contributed equally to the study (Zixu Zhao, Zejun Yang).

## Abstract

**Background:** Atrial fibrillation (AF) recurrence is common in patients after catheter ablation. Previous studies have demonstrated a lower risk of AF recurrence after ablation with the use of sodium-glucose cotransporter 2 inhibitors (SGLT2i) among patients with diabetes or heart failure. However, the effects of SGLT2i in AF patients after catheter ablation without current indications for SGLT2i were uncertain.

**Objective:** This trial aims to evaluate the effect of dapagliflozin on AF burden after ablation in persistent atrial fibrillation (PeAF) patients.

**Study Design:** Efficacy of DApagliflozin on REcurrence after catheter ablation for Atrial Fibrillation (DARE-AF) is a parallel-group, randomized, open-label randomized controlled trial. We aim to enroll 200 patients with de novo atrial fibrillation (AF) undergoing catheter ablation, and patients with class I indications for SGLT2 Inh (diabetes, heart failure, or chronic kidney disease) are excluded. PeAF patients will be randomized in a 1:1 ratio to intervention or control group. Patients in the intervention group will receive dapagliflozin 10mg once daily for three months. The primary outcome is AF burden accessed by 7-day single-lead electrocardiogram patches at three months after ablation.

**Conclusions:** DARE-AF is the first clinical trial, aiming to evaluate the effect of 3-month treatment with dapagliflozin on AF burden after catheter ablation in PeAF patients without current indications.

## 1 ADMINISTRATIVE INFORMATION

### 1.1 Study identifiers

- ClinicalTrial.gov register Identifier: NCT06433479, Date: 23 May 2024

### 1.2 Revision history

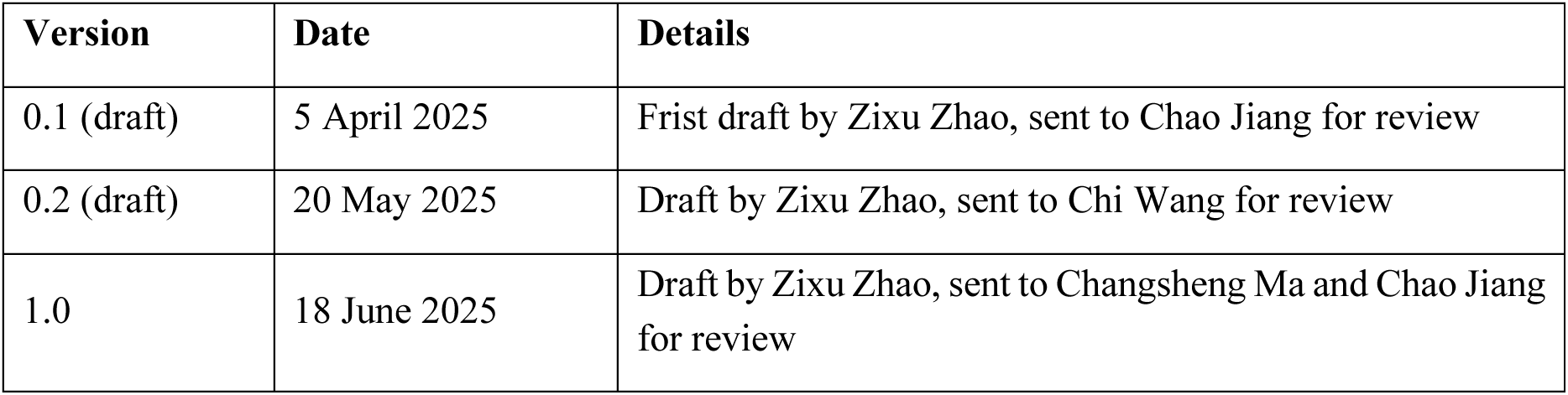

### 1.3 Contributors to the statistical analysis plan

Roles and responsibilities

1.3 Contributors to the statistical analysis plan Roles and responsibilities
| Name | Affiliation | Role on study | SAP contribution |
| --- | --- | --- | --- |
| Dr. Zixu Zhao | Beijing Anzhen Hospital, Capital Medical University | Study coordinator | Prepare initial draft and subsequent versions |
| Dr. Zejun Yang | Beijing Anzhen Hospital, Capital Medical University | Study coordinator | Reviewed every draft and approved final version |
| Mrs. Chi Wang | Heart Health Research Center, Beijing | Statistician | Prepare initial draft and subsequent versions |
| Prof. Chao Jiang | Beijing Anzhen Hospital, Capital Medical University | Principal investigator | Reviewed every draft and approved final version |
| Prof. Jianzeng Dong | Beijing Anzhen Hospital, Capital Medical University | Principal investigator | Reviewed every draft and approved final version |
| Prof. Changsheng Ma | Beijing Anzhen Hospital, Capital Medical University | Principal investigator | Reviewed every draft and approved final version |

### 1.4 Approvals

The undersigned have reviewed this plan and approve it as final. They find it to be consistent with the requirements of the protocol as it applies to their respective areas. They also find it to be compliant with International Conference on Harmonization (ICH-E9) principles and confirm that this analysis plan was developed in a completely blinded manner (i.e., without knowledge of the effect of the intervention[s] being assessed)

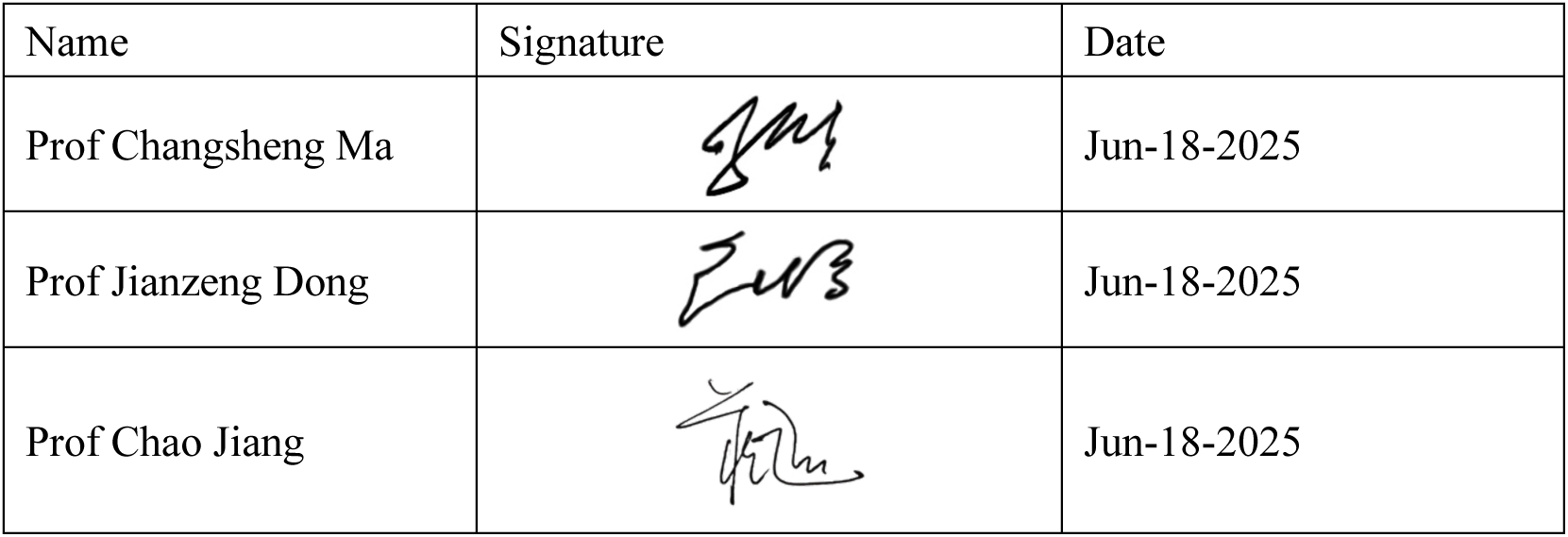

| Name | Signature | Date |
| --- | --- | --- |
| Prof Changsheng Ma |  | Jun-18-2025 |
| Prof Jianzeng Dong |  | Jun-18-2025 |
| Prof Chao Jiang |  | Jun-18-2025 |

## 2. INTRODUCTION

Atrial fibrillation (AF) is one of the most common arrhythmias, often linked to serious complications like stroke or heart failure (HF), resulting in a global socioeconomic burden.^1–3^ Catheter ablation as the first-line therapy for rhythm control can maintain sinus rhythm, reduce AF burden, and improve the patient’s quality of life.^4–7^ However, recurrence still occurs in 30-50% of patients after ablation, and there is currently no effective strategy to reduce AF recurrence after catheter ablation.^8,9^

Sodium-glucose cotransporter 2 inhibitors (SGLT2i) as an antihyperglycemic agent, have demonstrated efficiency in improving prognosis for patients with diabetes, HF and chronic kidney disease.^10,11^ A post hoc study demonstrated that dapagliflozin, an SGLT2i agent, might reduce AF or atrial flutter (AFL) events among patients with diabetes.^12^ Data from our previous cohort studies and meta-analysis demonstrated a lower risk of AF recurrence using SGLT2i among patients with diabetes and HF after catheter ablation.^13,14^ However, the beneficial effects of SGLT2i in AF patients after catheter ablation without current indications including diabetes, HF or chronic kidney disease were uncertain.

To test this hypothesis, our study aims to evaluate the effect of dapagliflozin on AF burden after ablation in persistent atrial fibrillation (PeAF) patients to determine the potential antiarrhythmic effect of SGLT2i in AF patients.

### 2.1 Rationale for this study (DARE-AF)

Efficacy of DApagliflozin on REcurrence after catheter ablation for Atrial Fibrillation (DARE-AF) is a prospective, open-label, parallel assignment randomized controlled trial (clinicaltrials.gov; NCT06433479). The study was reviewed and approved by the Beijing Anzhen Hospital ethics committee. After providing written informed consent, eligible patients are randomized 1:1 to dapagliflozin 10mg once daily for 3 months or a control group.

## 3 STUDY OBJECTIVES

### 3.1 Primary objective

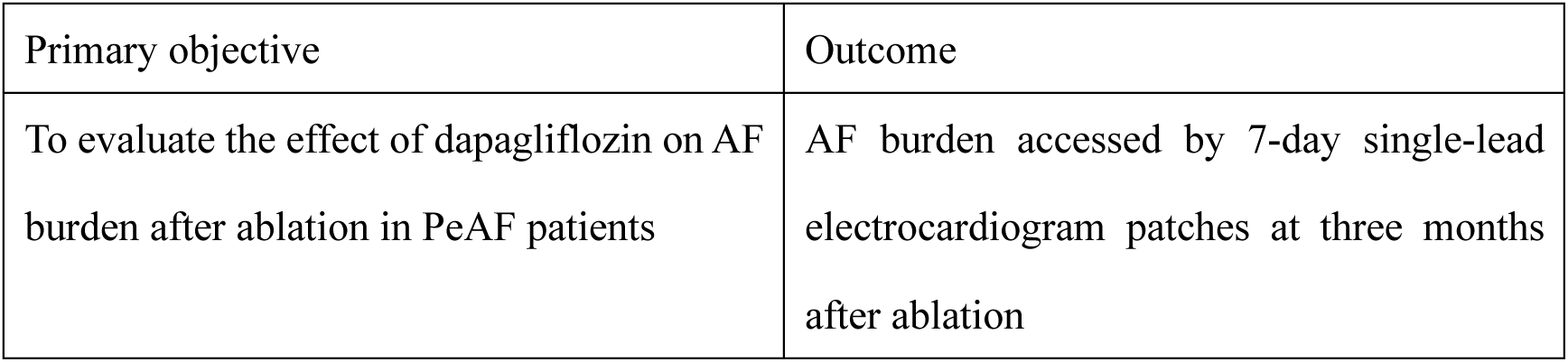

|  |  |
| --- | --- |
| Primary objective | Outcome |
| To evaluate the effect of dapagliflozin on AF burden after ablation in PeAF patients | AF burden assessed by 7-day single-lead electrocardiogram patches at three months after ablation |

### 3.2 Secondary objective

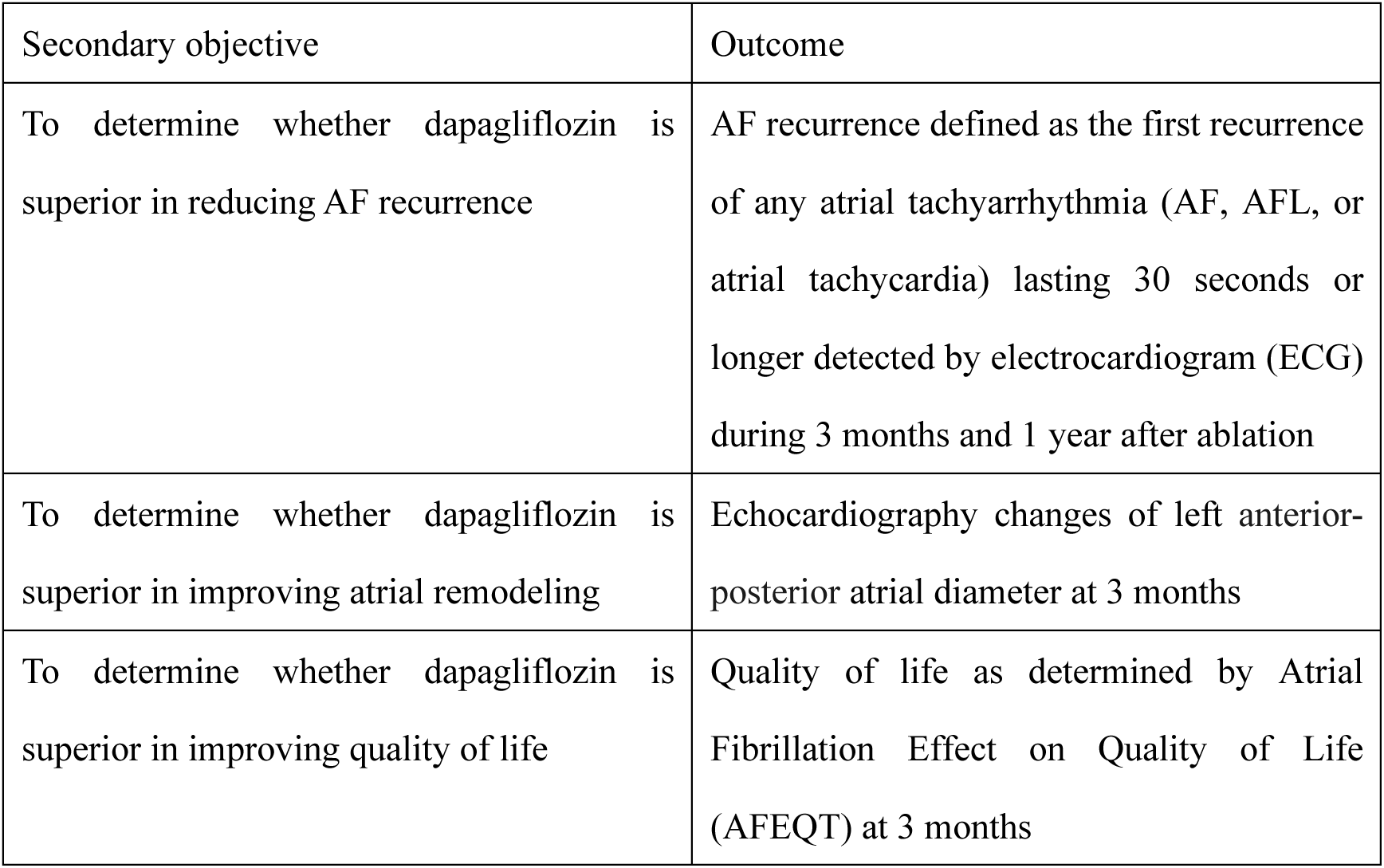

## 4 POPULATIONS TO BE ANALYZED

### 4.1 Intention-to-treat population (ITT)

The data analysis of the DARE-AF study will be performed by intention-to-treat analysis. Therefore, the primary endpoint will be analyzed in all patients with available primary outcome data according to their randomized treatment assignments, regardless of post-randomization medical care.

#### 4.2.1 Inclusion Criteria

1. age between 18-80 years
2. diagnosed with PeAF based on ECG or Holter
3. planned de novo catheter ablation for AF
4. have the capacity to understand and sign an informed consent form.

#### 4.2.2 Exclusion Criteria

1. diagnosed with PeAF longer than 5 years or left atrial anterior-posterior diameter ≥ 50mm
2. diagnosed with AF secondary to reversible causes (such as hyperthyroidism, acute infection, etc.)
3. severe structural heart disease (hypertrophic cardiomyopathy, rheumatic heart disease, dilated cardiomyopathy)
4. currently taking SGLT2i
5. with the following Class I indications for SGLT2i:

i. type 2 diabetes
ii. history of HF, including HF with reduced ejection fraction, mildly reduced ejection fraction, or preserved ejection fraction
iii. chronic kidney disease with eGFR 20-60 ml/min/1.73m^2^
6. with the following contraindications of SGLT2i:

i. previous allergic reactions to dapagliflozin
ii. end-stage renal failure or dialysis
iii. type 1 diabetes, or previous diabetic ketoacidosis
7. severe hypoglycemia or genitourinary infection in the past 12 months
8. hypovolemia or hypotension
9. planned surgery or other interventional procedure within 3 months
10. other arrhythmias mandating anti-arrhythmic drug therapy
11. intracardiac thrombus
12. active infection
13. unable to give informed consent
14. women of childbearing potential
15. currently enrolled in another clinical study
16. other conditions unsuitable to participate in this study judged by investigators

## 5 DEMOGRAPHIC AND BASELINE CHARACTERISTICS

Baseline characteristics (demographic, medications, echocardiography, if available) will be summarized overall and by treatment group for the ITT population. Continuous variables will be described as mean ± standard deviation (SD) or median (interquartile range), and categorical variables will be described as counts (percentages). Baseline characteristics of patients will be compared between the two groups using Student t-tests for normally distributed continuous variables, Wilcoxon rank sum tests for skewed continuous variables and Chi-Square tests for categorical variables.

## 6 COMPLIANCE

### 6.1 Visit adherence

Adherence to the visit schedule will be summarized by visit at 30 days, 60 days, and 90 days after randomization in the ITT population as follows:

- Frequency and percentage of participants having completed due visit
- The reason for not completing the visit

### 6.2 Medication adherence

After randomization, participants in the intervention group will receive pill containers with drug supply for the treatment period of 3 months. The first dose of dapagliflozin will be suggested to be taken 1 day after the procedures. The percentage of study drug compliance for the overall treatment period will be derived for each patient based on self-reported pill counts as the number of pills taken (dispensed - remained). The expected number of pills taken is defined as 1*(date of last dose - date of first dose + 1), excluding days of interruption. Full compliance with dapagliflozin is defined as self-reported intake of ≥80% of dapagliflozin.

Patients in the intervention group will have urine tests at least once within the 3-month follow-up. The medication adherence is assessed through the results of glycosuria. Notably, glycosuria in urine tests can be negative in participants without diabetes who received SGLT2i, as a previous study showed that 14.3% of healthy Chinese subjects had negative glycosuria after administration of dapagliflozin 10mg.^15^ Urine glucose secretion in response to SGLT2i could be variable across the population, probably related to the function and expression of the SGLT2 receptor in human renal. Therefore, overall medication compliance in the intervention group will be considered reasonable if 85% of patients have positive glycosuria during the 3-month follow-up. Besides, for patients with negative glycosuria, the follow-up staff will repeatedly confirm the data of medication time and remaining dose with the patient to ensure the patient’s medication compliance.

Medication adherence to dapagliflozin will be summarized as follows:

- Full compliance: frequency and percentage of participants who reported to have taken ≥80% of dapagliflozin
- Frequency and percentage of participants with drug continuation and discontinuation in 1-, 2-, and 3-month follow-up. Reasons for discontinuation will also be collected
- Frequency and percentage of participants with glycosuria positive for urine tests during the 3-month follow-up Use of oral anticoagulation and antiarrhythmic drugs will be described in both groups over time.

## 7 STUDY FOLLOW-UP

In-person or telephonic follow-up visits will be scheduled at 1-, 2-, 3-, 6-, and 12-month follow-up from the time of randomization.

### 7.1 Handling of non-compliance with study treatment or follow-up

A randomized subject who permanently discontinues dapagliflozin before the end of the treatment visit for any reason is defined as having had a permanent discontinuation of study medication (including subjects who will be randomized but never started taking any study medication). The reason for the permanent discontinuation of study medication will be collected. In this study, outcome events and clinical data will be collected until the end of the study, even if patients are no longer taking study medication. After the patients are randomized, the patients or their families and doctors will keep WeChat (a Chinese social networking software similar to Facebook) and phone numbers for each other to avoid a loss to follow-up. Every effort will be made to continue to follow the subject and survival status information and encourage patients’ compliance with drugs must be determined for all subjects at the end of the study.

In case of no contact, the participant will be censored on their last day of available contact during the study. Subjects who prematurely withdraw from treatment will not be withdrawn from the study and will be followed until the end of the study. Participants who withdrew consent or died will be censored on the date they withdrew consent or the date of death, respectively.

### 7.2 Handling of 7-day ECG patch data at three months

Patients are asked to wear the ECG monitor patch during a monitoring period accessed by 7-day single-lead electrocardiogram patches at three months after ablation. If the patient does not wear the patch for 72 consecutive hours, reminding message will be triggered. It is expected that some patients may have inadequate time to record the ECG patches to provide the end-of-study single-lead ECG assessment stipulated in the protocol. However, patients with less than 72-hour accumulative ECG monitoring will continue to be encouraged to monitor until the patients’ monitoring time is ≥72 hours unless the patient is lost to follow-up.

### 7.3 Handling of missing date information

Unless otherwise indicated in this section, missing values within follow-up will be treated as ‘missing’. No attempt will be made to use a computerized algorithm to impute missing post-randomization values and only observed values will be used for primary analysis.

When an event date is not known, the investigator will be asked to provide a best estimate as to when the event occurred. Even though the exact date of an event is unknown, the investigator often does know some information that will indicate the approximate date. This information can be meaningfully incorporated into the estimated date recorded. This estimated date should be the middle date within the period that the event is known to have occurred. If no information is known, then the date in the middle of the plausible time should be given based on the last contact with the patient before the event and the date of contact when information about the event is known.

## 8 OUTCOMES

### 8.1 Primary outcome

The primary outcome is the AF burden at 3 months after ablation. AF burden is defined as the proportion of time an individual is in any atrial tachyarrhythmia (AF, AFL, or atrial tachycardia) during a monitoring period accessed by 7-day single-lead ECG patches.

### 8.2 Secondary outcome

The secondary outcomes are (1) the proportion of participants with AF burden at 3 months after ablation <0.1%, 0.1-9.9%, and ≥10%; (2) the proportion of participants with AF episode duration ≥1 hour at 3 months after ablation; (3) AF recurrence during 3 months after ablation, defined as the first recurrence of any atrial tachyarrhythmia (AF, AFL, or atrial tachycardia) lasting 30 seconds or longer detected by ECG during 3 months after ablation; (4) echocardiography changes of left anterior-posterior atrial diameter at 3 months; (5) quality of life as determined by Atrial Fibrillation Effect on Quality of Life (AFEQT) questionnaire at 3 months.

### 8.3 Adverse events

An adverse event (AE) is the development of an undesirable medical condition or the deterioration of a pre-existing medical condition following or during exposure to a pharmaceutical product, whether or not considered causally related to the drug.

#### 8.3.1 Serious adverse events (SAE)

SAE is defined as any event that results in significant harm or increased risk for the subject or others (including individuals who are not research subjects). These include:

- Death
- Life-threatening adverse experience
- Inpatient hospitalization or prolongation of existing hospitalization
- Persistent or significant disability/incapacity
- Congenital anomaly/birth defect
- Intervention to prevent permanent impairment or damage

#### 8.3.2 Adverse event of special interest (AESI)

AESI is defined as expected adverse events associated with the use of dapagliflozin in the intervention group. AESI in DARE-AF included:

- Hypotension: hypotension events are defined as events that occur in patients with clinically relevant symptoms or signs of suspected hypotension (dizziness, syncope), and confirmed and sustained blood pressure <90/60 mmHg.
- Hypoglycemia: Major hypoglycemic event defined as symptomatic events requiring external assistance due to severe impairment in consciousness or behavior, and prompt recovery after glucose or glucagon administration. Plasma glucose measurements may not be available during such an event, but neurological recovery attributable to the restoration of plasma glucose to normal is considered sufficient evidence that a low blood glucose concentration induced the event.
- Urinary tract or genital infection: The diagnosis of vaginitis, vulvovaginitis, vulvitis or balanitis can be made basedon physical examinations, culture of secretions or a therapeutic response to treatment of fungal or other vaginal pathogens. Clinical judgment and local standards of care should apply to decisions concerning therapy.
- Diabetic ketoacidosis: Patients treated with dapagliflozin who present with signs and symptoms consistent with ketoacidosis, including nausea, vomiting, abdominal pain, malaise, and shortness of breath, should be assessed for ketoacidosis, even if blood glucose levels are below 14 mmol/L (250 mg/dL). The diagnosis of ketoacidosis in these patients can be based on arterial pH ≤7.30, serum bicarbonate levels <15 mmol/L and measurement of serum beta-hydroxybutrate levels. Other diagnostic criteria that can support the diagnosis of ketoacidosis are urine ketones and anion gap >10 mmol/L.
- Renal dysfunction: occurrence of any of the components of this composite: 1. confirmed sustained decrease in eGFR to eGFR <60 mL/min/1.73m^2^ or ≥50% sustained decline in eGFR from baseline using the CKD-EPI equation; 2. dialysis treatment or receiving a renal transplant

## 9 GENERAL STATISTICAL CONSIDERATIONS

### 9.1 Sample size

According to the data of previous research,^16–19^ the AF burden of the control group is expected to be 10% at 3 months after ablation, and the AF burden of the dapagliflozin group is 30% lower, which is expected to be 7%. With a standard deviation of 6% and a bilateral α value of 0.05, 172 patients will provide a power of 90% for the primary outcome. A total of 200 patients (100 in each group) are intended to be enrolled based on the loss of follow-up rate of 5%.

### 9.2 Primary Outcome

Descriptive statistics will be used for the primary outcomes based on the ITT population. The AF burden will be described with Mean ± SD. The AF burden of the dapagliflozin group versus the control group will be compared using the stratified Mann-Whitney U test with randomization factors of baseline BMI (<24kg/m^2^ vs. ≥24kg/m^2^) and baseline left atrial anterior-posterior diameter (<45mm vs. ≥45mm) as strata. A 2-sided P value of <0.05 will be considered statistically significant for statistical analyses.

The primary outcome will also be evaluated with linear regression as a sensitivity test. Based on previous research,^20^ the linear regression will be used to assess the log(e) AF burden as an independent variable. The log(e) AF burden is back-transformed and presented as a geometric mean. The stratification factors of baseline BMI (<24kg/m^2^ vs. ≥24kg/m^2^) and baseline left atrial anterior-posterior diameter (<45mm vs. ≥45mm) will be included.

### 9.3 Secondary Outcomes

All secondary outcome analyses will be performed in the ITT population.

#### 9.3.1 AF burden divided by proportions

According to current literature and expert consensus,^21–23^ an AF burden exceeding 0.1% is considered clinically significant. Therefore, AF burden will be categorized into three groups: <0.1%, 0.1-9.9%, and ≥10%. The distribution of patients across these AF burden strata will be compared between groups using the chi-square test.

#### 9.3.2 AF episode durations ≥1 h divided by proportions

According to current literature and expert consensus,^21–23^ an AF episode duration exceeding 1 hour is considered clinically significant. We defined a binary outcome as the occurrence of at least one AF episode lasting≥1 hour during follow-up. Logistic regression is performed to compare the odds ratio of this outcome between groups. Results are presented as odds ratios (ORs) with corresponding 95% confidence intervals (CIs).

#### 9.3.3 AF recurrence

Time to AF recurrence is analyzed using Cox proportional hazard models between the dapagliflozin and control groups during 3-month follow-ups. The effect of dapagliflozin relative to control will be presented as HR and its 95% CI. The cumulative incidence of AF recurrence will be estimated using the Kaplan–Meier method based on time to first event.

#### 9.3.4 Changes of left atrial anterior–posterior diameter

The change in left atrial anterior–posterior diameter from baseline to the end of follow-up will be compared between the two groups using the analysis of covariance (ANCOVA) model with baseline left atrial anterior-posterior diameter as a covariate.

#### 9.3.5 Changes of AFEQT

The change in AFEQT from baseline to the end of follow-up will be compared between the two groups using the ANCOVA model with baseline AFEQT as a covariate.

### 9.4 Sensitivity analysis of the primary outcome

Sensitivity analysis will be conducted in patients with positive glucosuria in urine tests during follow-up. Negative glycosuria may partially reflect suboptimal medication adherence in patients. Besides, sensitivity analysis will be performed in patients with full compliance, defined as those who reported having taken ≥80% of dapagliflozin. We will also perform a sensitivity analysis in patients with ≥72-hour cumulative ECG monitoring accessed by 7-day single-lead ECG patches at 3 months after ablation.

### 9.5 Safety analysis

SAE or AESI will be collected from the time of enrollment throughout the treatment period. SAE or AESI will be recorded from the time of signing of the informed consent form. If the investigator becomes aware of an SAE or AESI with a suspected causal relationship to the investigational medicinal product that occurs after the end of the clinical study in a participant treated by him or her, the investigator shall, without undue delay, report the SAE or AESI. At each contact with the subject, the study team must seek information on adverse events by specific questioning and, as appropriate, by examination. Information on all adverse events should be recorded immediately in the source document and also in the appropriate adverse event section of the case report form. All related signs, symptoms, and abnormal diagnostic, laboratory or procedure results should be recorded.

All adverse events occurring during the study period must be recorded. The clinical course of each event should be followed until resolution, stabilization, or until it has been ultimately determined that the study treatment or participation is not the probable cause. SAEs or AESIs that are still ongoing at the end of the study period must be followed up, to determine the final outcome.

Information for patient-reported SAE and AESI for all randomized participants is specified in section 8.3 of the protocol. Summary statistics will be generated for SAE or AESI occurring on or after randomization, overall and according to randomized treatment. A summary table of the total number and percent of patients with SAE or AESI leading to temporary interruption, amputations, and preceding events per treatment group will be provided.

## 10 SUBGROUP ANALYSIS

The primary outcome AF burden at 3 months after catheter ablation will be investigated in prespecified subgroups. The p-value of the interaction term between the treatment and subgroups will be reported.

1. Sex (male, female);
2. Age (<60, ≥60);
3. AF duration (<1 year, 1-5 year);
4. Body mass index (<24kg/m^2^, ≥24kg/m^2^);
5. left anterior-posterior atrial diameter (<45mm, ≥45mm);
6. CHA_2_DS_2_-VA score (<2, ≥2);

The analysis for each subgroup will be performed by first stratifying the ITT population according to each subgroup variable. If the number of patients within a subgroup is too small, e.g. less than 5% of the overall population, the analyses may not be performed, or the subgroup levels may be combined. The results will be displayed on a forest plot, including the point estimate and 95% confidence interval.

## Data Availability

All data produced in the present work are contained in the manuscript

## 12. APPENDIX 1: PROPOSED TABLES AND FIGURES

**Table 1:**
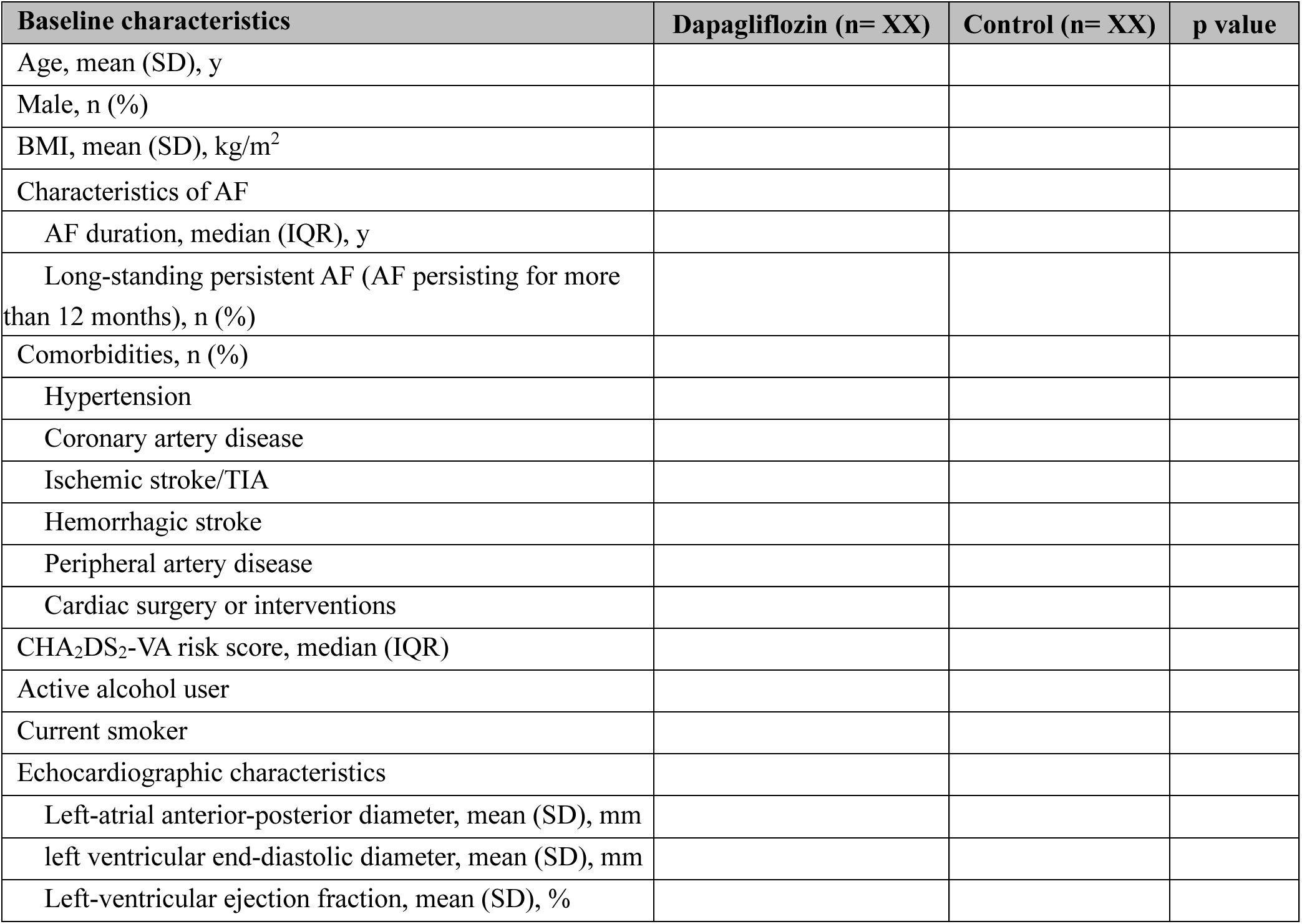
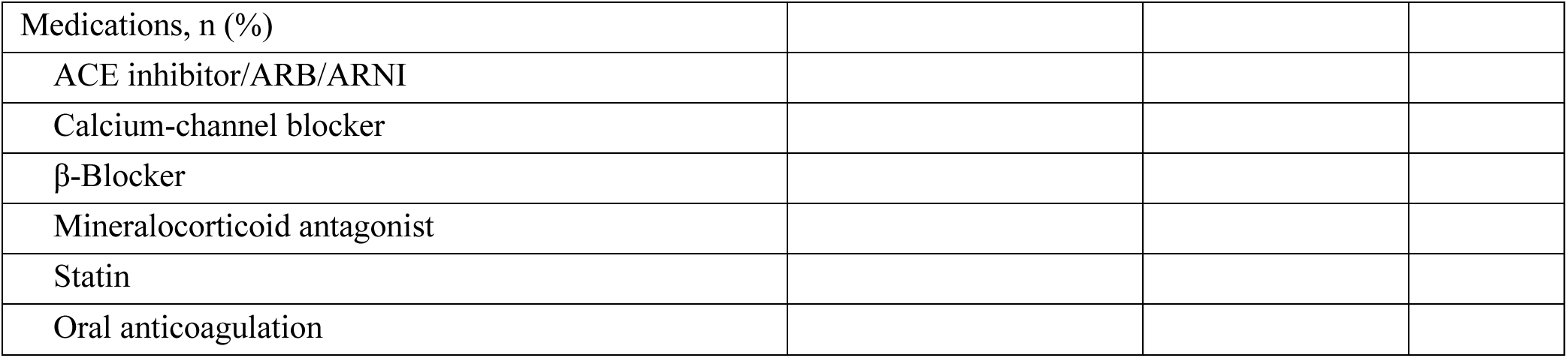
Baseline characteristics.

| Baseline characteristics | Dapagliflozin (n= XX) | Control (n= XX) | p value |
| --- | --- | --- | --- |
| Age, mean (SD), y |  |  |  |
| Male, n (%) |  |  |  |
| BMI, mean (SD), kg/m<sup>2</sup> |  |  |  |
| Characteristics of AF |  |  |  |
| AF duration, median (IQR), y |  |  |  |
| Long-standing persistent AF (AF persisting for more than 12 months), n (%) |  |  |  |
| Comorbidities, n (%) |  |  |  |
| Hypertension |  |  |  |
| Coronary artery disease |  |  |  |
| Ischemic stroke/TIA |  |  |  |
| Hemorrhagic stroke |  |  |  |
| Peripheral artery disease |  |  |  |
| Cardiac surgery or interventions |  |  |  |
| CHA<sub>2</sub>DS<sub>2</sub>-VA risk score, median (IQR) |  |  |  |
| Active alcohol user |  |  |  |
| Current smoker |  |  |  |
| Echocardiographic characteristics |  |  |  |
| Left-atrial anterior-posterior diameter, mean (SD), mm |  |  |  |
| left ventricular end-diastolic diameter, mean (SD), mm |  |  |  |
| Left-ventricular ejection fraction, mean (SD), % |  |  |  |
| Medications, n (%) |  |  |  |
| ACE inhibitor/ARB/ARNI |  |  |  |
| Calcium-channel blocker |  |  |  |
| β-Blocker | | | |
| Mineralocorticoid antagonist |  |  |  |
| Statin |  |  |  |
| Oral anticoagulation |  |  |  |

**Table 2:** Primary and Secondary Outcomes.

| Outcomes | Dapagliflozin group<br>(n = XX) | Control group<br>(n = XX) | Estimate<br>(95% CI) | P value |
| --- | --- | --- | --- | --- |
| **Primary Outcomes** |  |  |  |  |
| AF burden at 3 months, mean (SD), %* |  |  |  |  |
| AF burden at 3 months, geometric mean difference** | / | / |  |  |
| **Secondary outcomes** |  |  |  |  |
| AF burden at 3 months divided by proportions, n (%) |  |  |  |  |
| <0.1% |  |  |  |  |
| 0.1-9.9% |  |  |  |  |
| $\geq 10\%$ | | | | |
| Proportions of patients with AF episode durations $\geq 1$ h, n (%) | | | | |
| AF recurrence during 3 months, n (%) |  |  |  |  |
| Changes of LAD from baseline to 3 months, mean (SD), mm |  |  |  |  |
| Changes of AFEQT from baseline to 3 months, mean (SD) |  |  |  |  |

\* Using the stratified Mann-Whitney U test with randomization factors of baseline BMI (<24kg/m<sup>2</sup> vs. $\geq 24$ kg/m<sup>2</sup>) and baseline left atrial anterior-posterior diameter (<45mm vs. $\geq 45$ mm) as strata.
\*\* Using linear regression included the log(e) AF burden as a sensitivity test for the primary outcome.

**Figure 1:**
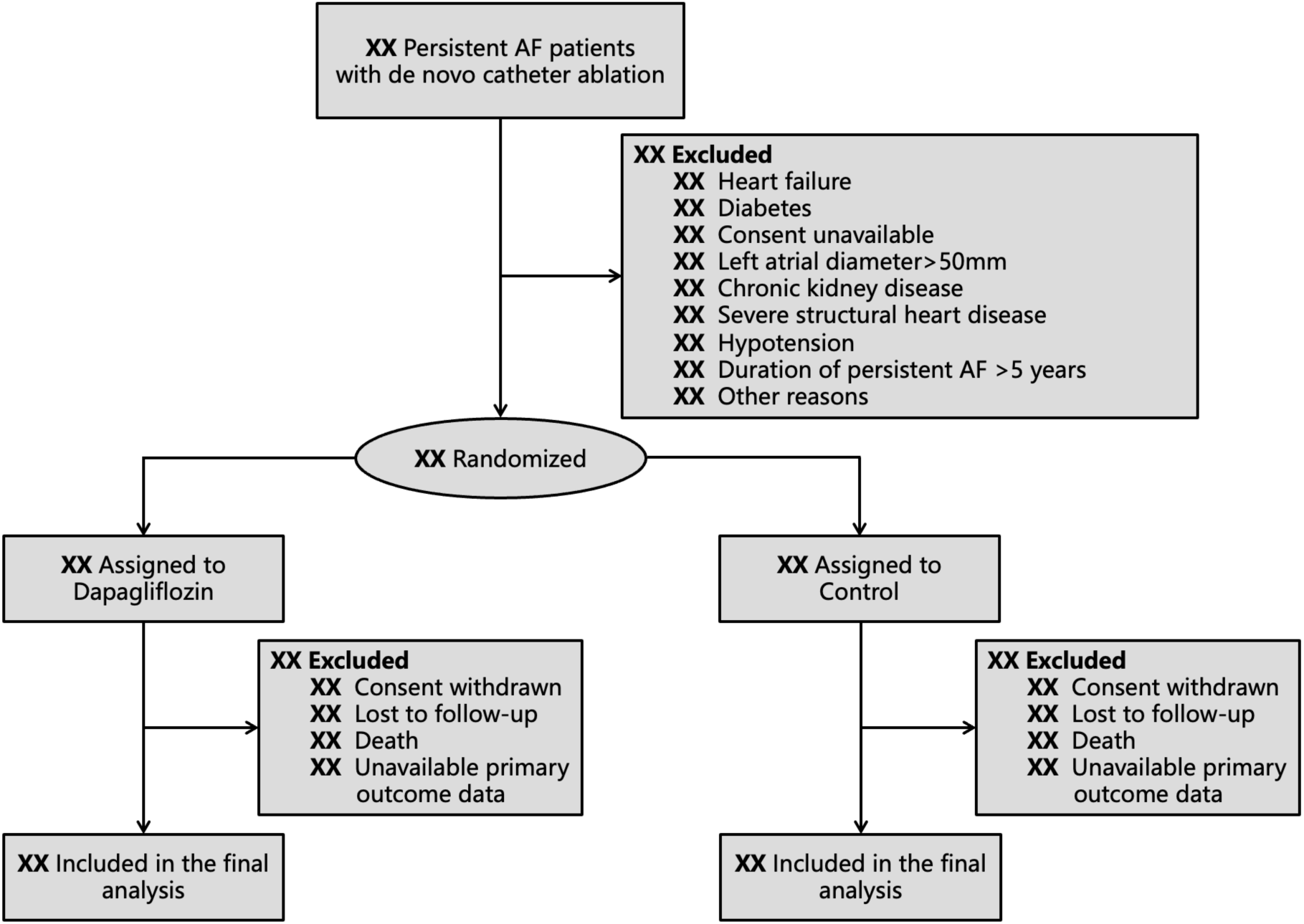
**The flow diagram**

**Figure 2:**
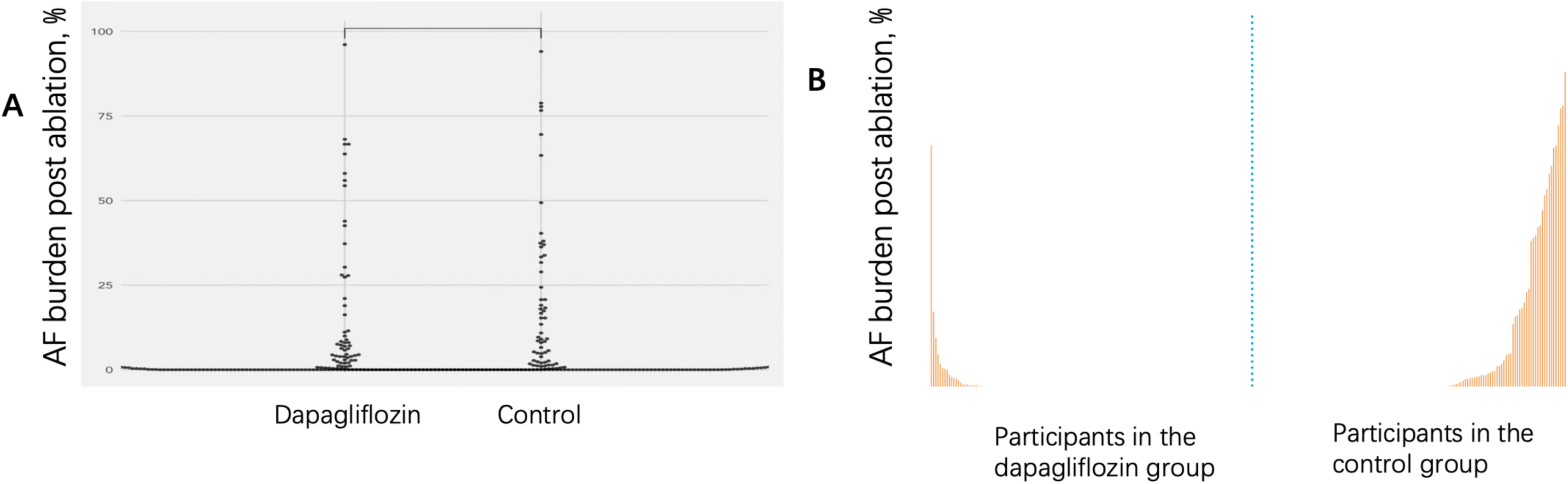
**AF burden at 3 months post-ablation between the groups. A. AF burden at 3 months post-ablation in control vs dapagliflozin group. B. Waterfall plot of AF burden at 3 months**

**Figure 3:**
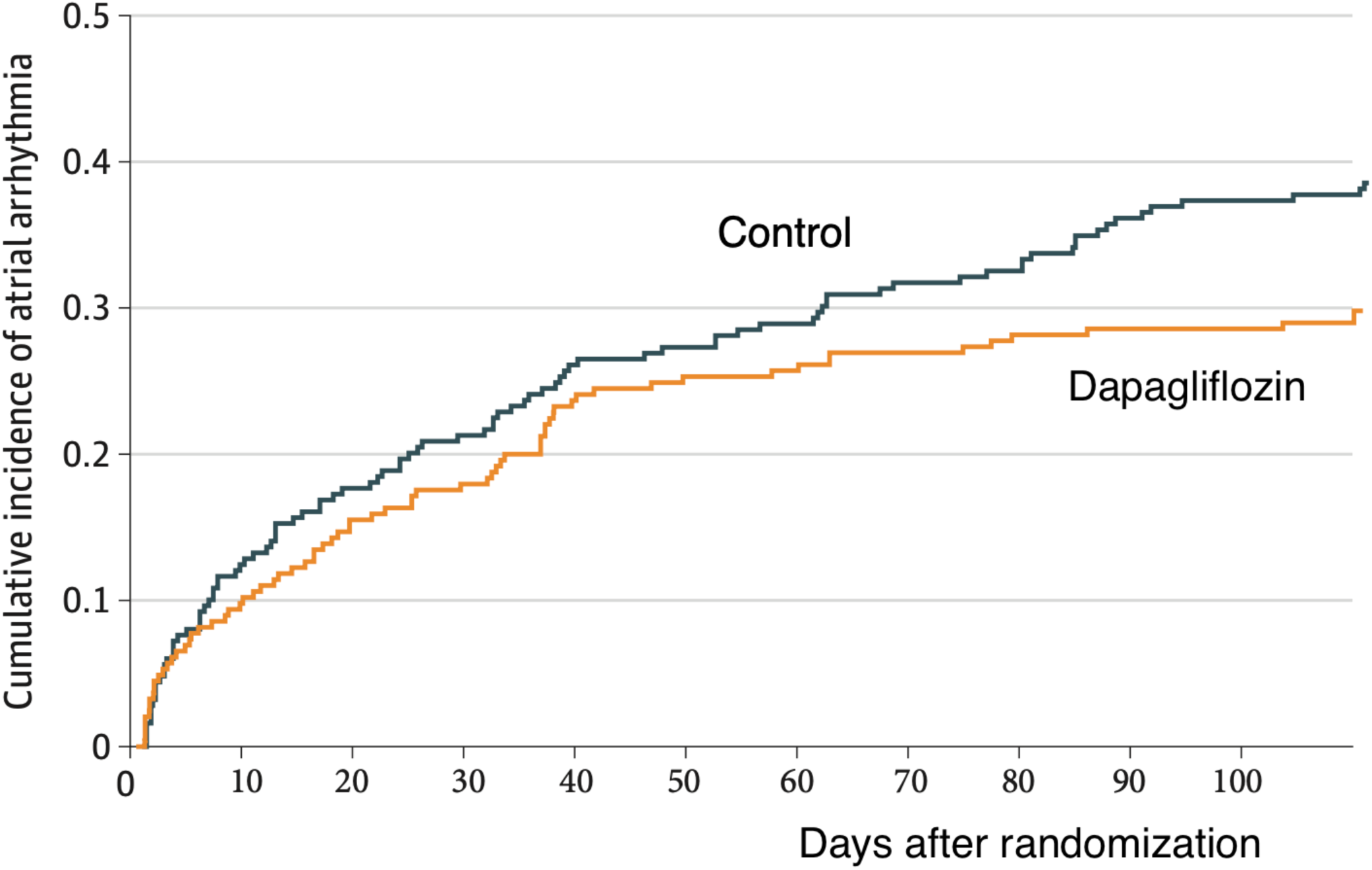
**Freedom from recurrence of atrial arrhythmias between two groups**

**Figure 4:**
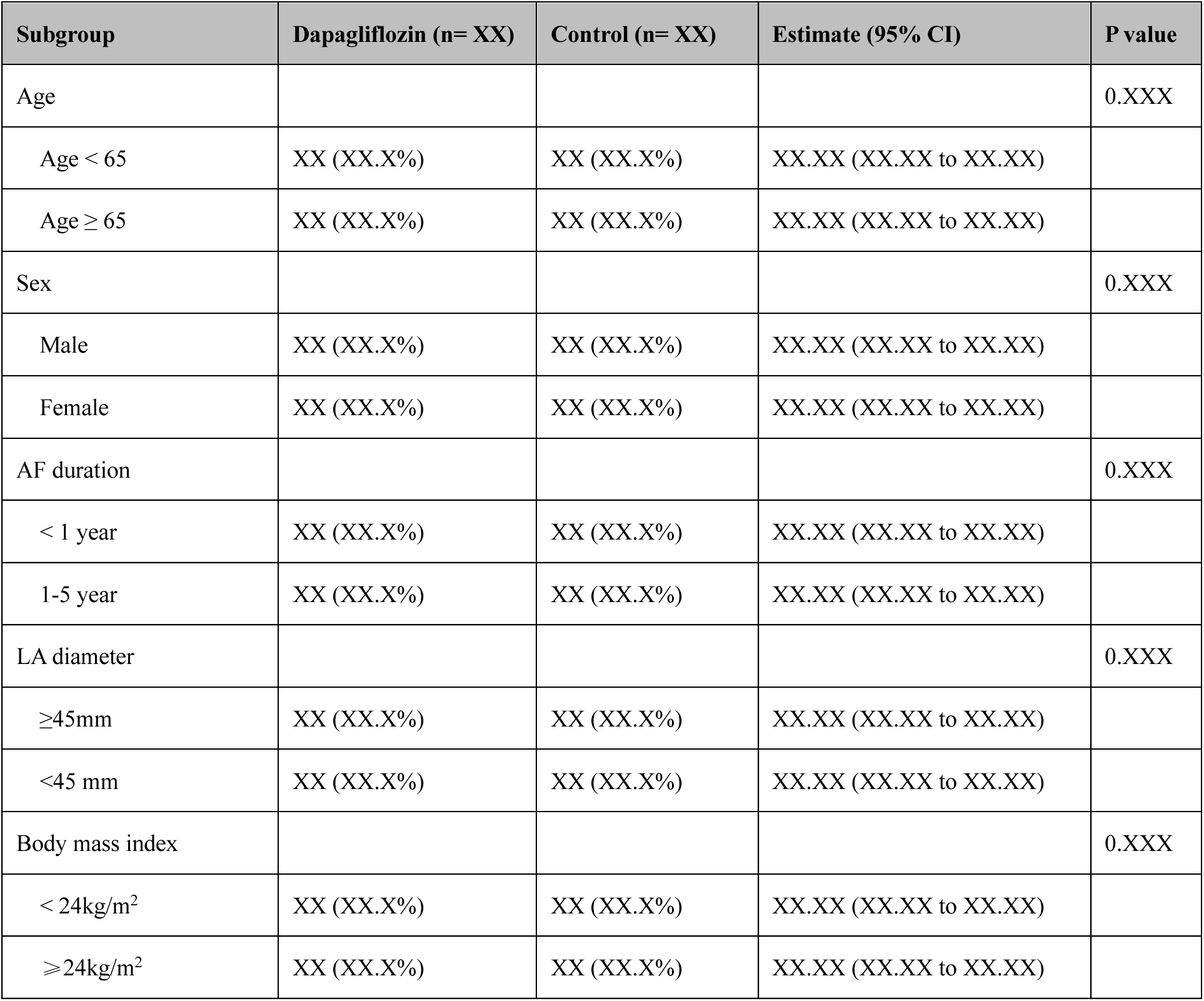
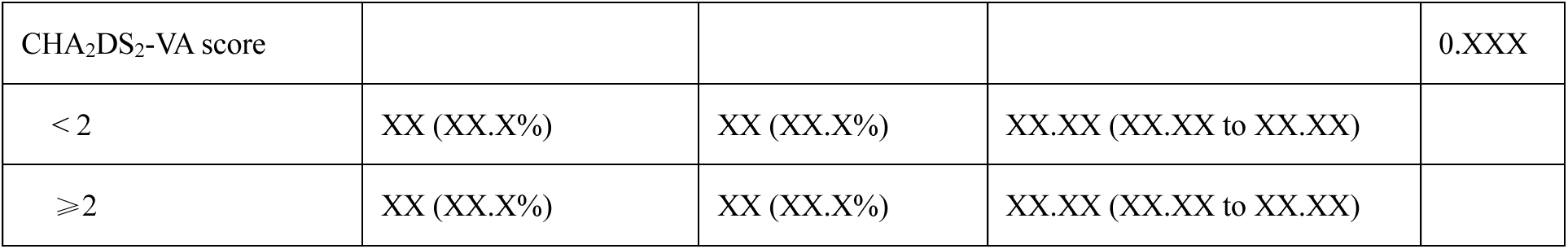
**subgroup study for the primary outcome**

## 13. APPENDIX 2: SUPPLEMENTARY MATERIALS

**Supplementary Table 1:** Procedural details.

| Procedural details | Dapagliflozin (n= XX) | Control (n= XX) | P value |
| --- | --- | --- | --- |
| Radiofrequency ablation, n |  |  |  |
| Pulsed Field Ablation, n |  |  |  |
| Total procedure duration, min; median (IQR) |  |  |  |
| Concomitant ablation, n |  |  |  |
| None |  |  |  |
| Cavotricuspid isthmus |  |  |  |
| Mitral isthmus |  |  |  |
| Roofline |  |  |  |
| Coronary sinus |  |  |  |
| Ethanol infusion via vein of Marshall |  |  |  |
| Others |  |  |  |
| Electrical cardioversion during procedure |  |  |  |

**Supplementary Table 2:**
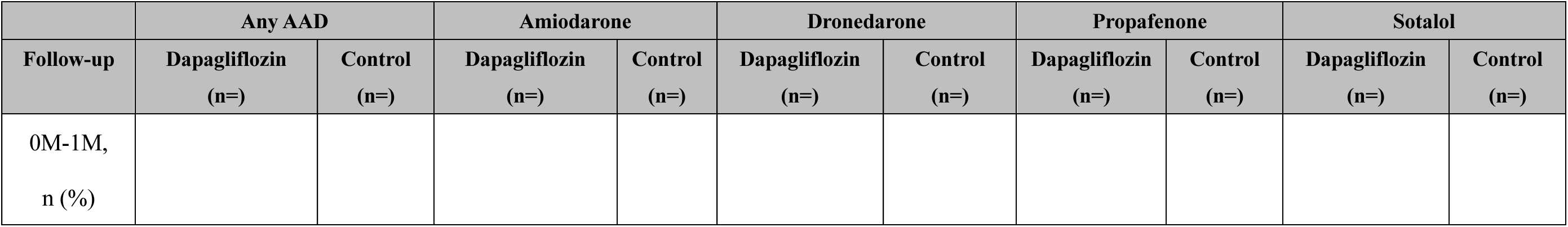
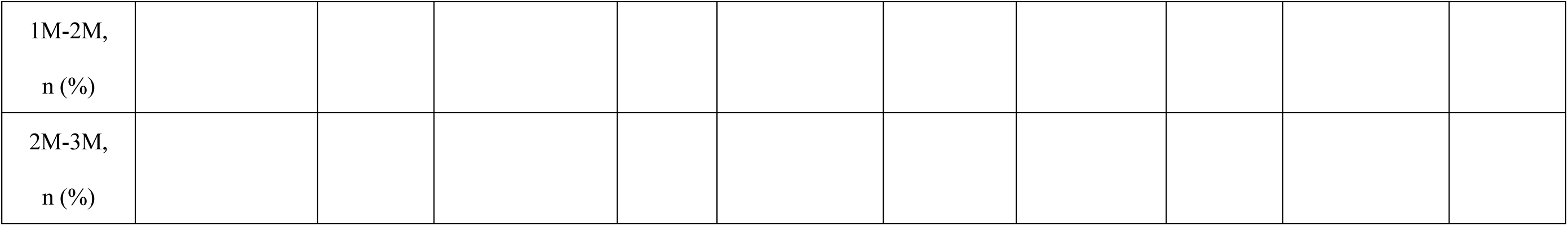
Use of antiarrhythmic drugs during follow-up.

| Follow-up | Any AAD |  | Amiodarone |  | Dronedarone |  | Propafenone |  | Sotalol |  |
| --- | --- | --- | --- | --- | --- | --- | --- | --- | --- | --- |
|  | Dapagliflozin<br>(n=) | Control<br>(n=) | Dapagliflozin<br>(n=) | Control<br>(n=) | Dapagliflozin<br>(n=) | Control<br>(n=) | Dapagliflozin<br>(n=) | Control<br>(n=) | Dapagliflozin<br>(n=) | Control<br>(n=) |
| 0M-1M,<br>n (%) |  |  |  |  |  |  |  |  |  |  |

|  |
| --- |
| 1M-2M,<br>n (%) |
| 2M-3M,<br>n (%) |

**Supplementary Table 3:** Adherence of dapagliflozin in the intervention group during follow-up.

| **Compliance of dapagliflozin** | **N** |
| --- | --- |
| **Full compliance 0M-3M (intake of $\geq 80\%$ dapagliflozin)** |  |
| **0M-1M drug compliance, n (%)** |  |
| Drug continuation |  |
| Drug discontinuation |  |
| Reasons for discontinuation |  |
| Hypotension |  |
| Urinary tract or genital infection |  |
| Other |  |
| **1M-2M drug compliance, n (%)** |  |
| Drug continuation |  |
| Drug discontinuation |  |
| Reasons for discontinuation |  |
| Hypotension |  |
| Urinary tract or genital infection |  |
| Other |  |

STATISTICAL ANALYSIS PLAN
| 2M-3M drug compliance, n (%) |
| --- |
| Drug continuation |
| Drug discontinuation |
| Reasons for discontinuation |
| Hypotension |
| Urinary tract or genital infection |
| Other |
| Urine tests during 3M follow-up |
| Glycosuria (-) |
| Glycosuria (+) |
| Glycosuria (++ - ++++) |

**Supplementary Table 4:** Adverse event.

| Adverse events | Dapagliflozin (n= XX) | Control (n= XX) |
| --- | --- | --- |
| Serious adverse events (SAE) |  |  |
| All-cause mortality |  |  |
| Cardiovascular hospitalization |  |  |
| Adverse events of special interest (AESI) |  |  |
| Hypotension |  |  |
| Hypoglycemia |  |  |
| Urinary tract or genital infection |  |  |
| Diabetic ketoacidosis |  |  |
| Renal dysfunction |  |  |

